# Factors Influencing The First and Second Peak of COVID-19 Global Cases: A Survival Analysis

**DOI:** 10.1101/2021.09.13.21263497

**Authors:** Yesar Ahmed Oshan, Begum Zainab, Dipankar Bandyopadhyay, Md Hasinur Rahaman Khan

## Abstract

**Objectives:** The number of reported cases continues to increase everyday, since the first case of COVID-19 was detected in Wuhan, China in December 2019. Using the global COVID-19 data of 188 countries extracted from the Our World in Data between January 22, 2020–January 18, 2021, this study attempts to explore the potential determinants of the number of days to reach the *first* and *second* peak of COVID-19 cases for all 188 countries.

**Methods:** A semi-parametric Cox proportional hazard (PH) model has been used to explore the covariates that are associated with the number of days to reach the *first* and *second* peak of global COVID-19 cases.

**Results:** As of January 18, 2021, the first and second peak were found in 175 and 59 countries, out of 188 countries, respectively. The median number of days to hit the first peak was 60 days for countries which have median age above 40 while the median number of days to hit the second peak was 267 days for countries which have population density above 500 per square kilometer. Countries having population density between 250 and 500 were 2.25 times more likely to experience the first peak of COVID-19 cases (95% CI: 1.15-4.45, *P* < 0.05) than countries which have population density below 25. Countries having population density between 100 and 250 were 67% less likely to get the second peak (95% CI: 0.119-0.908, *P* < 0.05) compared to countries which have population density below 25. Countries having cardiovascular death rates above 350 were 2.94 times more likely to get the first peak (95% CI: 1.59-5.43, *P* < 0.001). In contrast, countries having diabetes prevalence rate 3 to 12 were 85% less likely to experience the second peak of COVID-19 cases (95% CI: 0.036-0.680, *P* < 0.05) than countries which have diabetes prevalence rate below 3. Besides, highly significant difference is found in the Kaplan-Meier plots of the number of days to reach both peaks across different categories of the country’s Human Development Index.

**Conclusions:** The number of days to the first peak was considerably small in Asian & European countries but that to the second peak in the countries where diabetes prevalence was very higher. Country’s life expectancy had a significant effect on determining the first peak and so was the case for two other variables–the cardiovascular death rate and hospital beds per thousand. A contrast result was found for Human Development Index factor under the second peak. Additionally, it was found that the second peak was more likely to occur in more densely populated countries.

## 1 Introduction

Coronavirus disease (COVID-19) is an infectious disease caused by a newly discovered coronavirus. Most people infected with the COVID-19 virus will experience mild to moderate respiratory illness, and recover without requiring special treatment. Older people and those with underlying medical comorbidities, such as cardiovascular disease, diabetes, chronic respiratory disease and cancer are more likely to develop serious illness (Organization, 2020). The first human cases of COVID-19, the disease caused by the novel coronavirus causing COVID-19, subsequently named SARS-CoV-2, were first reported by officials in Wuhan City, China, in December 2019. Retrospective investigations by Chinese authorities have identified human cases with onset of symptoms, as early as December 2019 (Organization et al., 2020).

COVID-19 across many countries is getting severe day by day. By 18 February 2021, the total number of confirmed cases has reached to 110 million (Roser et al., 2021). This pandemic has extremely negative effects on human life and their daily activities. Public health, food systems, and the world economy are facing an immense challenge. The social catastrophe triggered by the epidemic is devastating. According to WHO, 150 million people are at risk of falling into extreme poverty by 2021.

Health care delivery organizations are facing unforeseen challenges due to COVID-19. For instance, hospitals lack adequate capacity to manage this rising number of patient admissions. A large number of patients is dying due to lack of intensive care unit (ICU) beds and ventilators at the proper time. Besides, safety measures are inadequate for healthcare workers. Knowing the peaks of COVID-19 cases is very essential for many reasons including health care delivery departments and organizations. Based on an informed estimate of the peaks and associated covariates, they can make enough preparations to fight adequately against this invisible enemy. Many countries have already experienced the first and second waves of COVID-19. By January 2021, the first peak of COVID-19 had been found almost in all the countries, and the second peak had been found in a number of countries.

A number of studies has been conducted with survival analysis techniques to COVID-19 data. For example, Leulseged et al. (2020) applied survival analysis to estimate time to getting off supplemental oxygen therapy, and identify predictors among COVID-19 patients admitted to Millennium COVID-19 Care Center in Addis Ababa, Ethiopia. Also, Oulhaj et al. (2020) investigated the bias (through extensive simulations) in estimating the hazard ratio (HR) and the absolute risk reduction (ARR) of death when competing risks are ignored. Furthermore, Thai et al. (2020), Salinas-Escudero et al. (2020) and Kundu and Mandal (2020) employed survival analysis techniques to evaluate COVID-19 risk assessments in Vietnamese, Mexican and Indian populations, respectively. In the realms of exploring machine learning techniques, Atlam et al. (2021) proposed a combination of autoencoder deep neural network and Cox regression to enhance prediction accuracy. However, none of the above studies developed a credible regression framework to investigate the hazards of various underlying factors, such as life expectancy, diabetes prevalence, etc, on the time-to-events of reaching the peaks.

Atlam et al. (2021) presented two systems Cox COVID-19 and Deep Cox COVID-19 that are based on Cox regression to study the survival analysis for COVID-19 and help hospitals to choose patients with better chances of survival and predict the most important symptoms (features) affecting survival probability. Salinas-Escudero et al. (2020) applied survival analysis to investigate the impact of COVID-19 on the Mexican population and found that the risk of dying at any time during follow-up was clearly higher for men, individuals in older age groups, people with chronic kidney disease and people hospitalized in public health services. Implementing survival analysis Castelnuovo et al. (2020) found that impaired renal function, elevated C-reactive protein and advanced age were major predictors of in-hospital death in a large cohort of unselected patients with COVID-19, admitted to 30 different clinical centres all over Italy.

Kundu and Mandal (2020) carried out a survival analysis to establish the variability in survivorship among age groups and sex at different levels that is national, state and district level in india. Ali et al. (2020) implemented survival analysis to determine risk factors for death in patients with COVID-19 admitted to the main public sector hospital in Somalia and identify interventions contributing to improved clinical outcome in a low-resource and fragile setting. Implementing multivariate Cox-PH model (Wang et al., 2020) found that preparation control measures of COVID-19 should involve the allocation of sufficient medical resources, especially in areas with older vulnerable populations and in areas that lack basic medical resources.

In this study, we primarily focused on evaluating the *time to hitting the first, and second peak* of global COVID-19 cases. Besides, we also attempted to determine the potential determinants that affect the peak time. The peak time of COVID-19 cases for each country was defined as time interval between the first case identification date and the peak hitting date. Since the peak time was not available for all the countries, we had incomplete peak time that demands to conduct survival analysis (Bewick et al., 2004) for failure time that is the time to hitting peak of COVID-19 cases by the country in our analysis.

## 2 Data and Variables

The COVID-19 dataset utilized in the analysis had been collected for 188 countries from *Our World in Data* (Roser et al., 2021) data repository site for the period January 22, 2020 to January 18, 2021. The dataset included country-level daily data on confirmed cases, deaths, testing, and other variables of potential interest. This data was assimilated with information brought from different sources, including the European Centre for Disease Prevention and Control, the International Organisation for Standardisation (ISO), national government reports, the Department of Economic and Social Affairs of the United Nations (UN), UN Population Division, UN Statistics Division, Oxford COVID-19 Government Response Tracker, the World Bank, the Global Burden of Disease Collaborative Network, and Eurostat of the Organisation for Economic Cooperation and Development.

To determine whether countries hit the first peak, both first and second peak, or no peaks, we produced country-wise bar plots with the date stated on the *x*-axis, and the number of daily COVID-19 cases stated on the *y*-axis. From bar plot of a country’s COVID-19 cases, we decided whether the country hits first peak, both first and second peak, or no peak at all. First or second peak is assumed to take place if after happening any peak the declining trend is observed for at least 2 weeks. But for second peak to happen we further observed whether the COVID-19 cases were in minimum level for at least two weeks. For selecting the peaks we consider Figure 1 as the benchmark.

**Figure 1:**
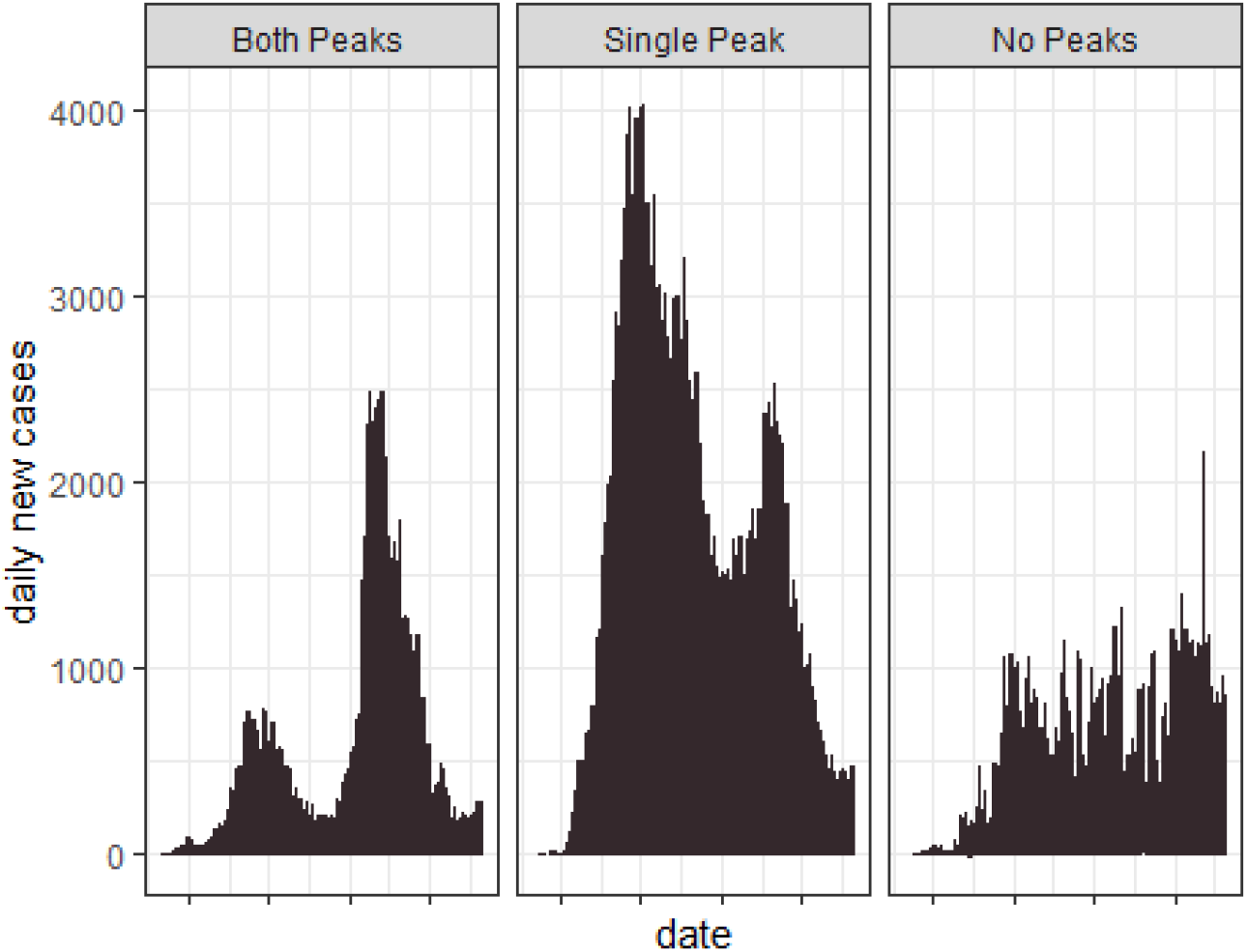
Defining and Understanding Peak

Subsequently, we extracted necessary variables for implementing to survival models and created two separate and independent datasets, corresponding to attaining the first and the second peak. A total of 188 countries were included. Focusing on the first peak dataset, countries that didn’t attend their first peak by the end of data collection date (January 21, 2021) were considered right-censored. Among the 188 countries, first peak was observed for 175 countries and the second peak was observed for 58 countries. According to this same data-driven procedure we found that third peak ocurred only for Luxembourg. Hence, we did not consider any third peak in this study. The ‘time’ and ‘status’ under the first peak were calculated as below.

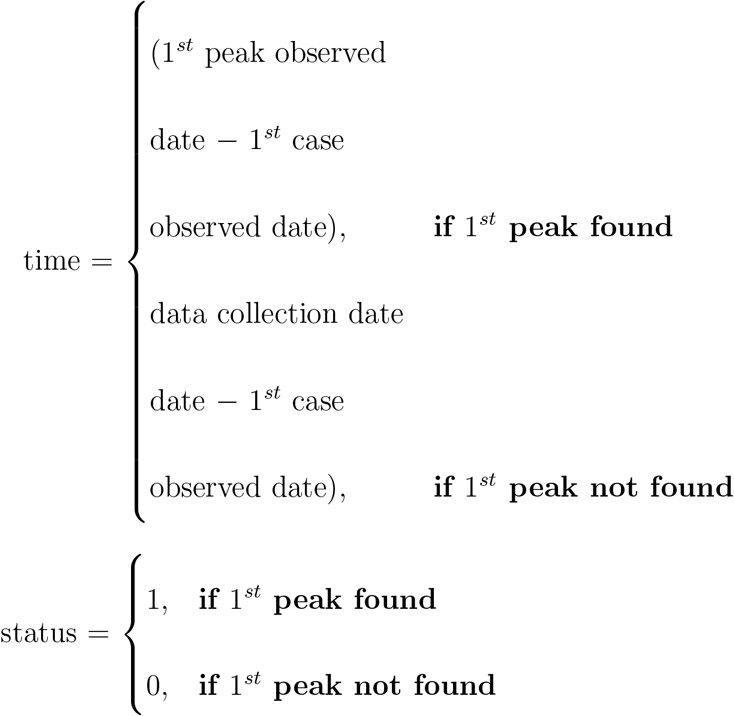

The ‘time’ and ‘status’ under the second peak were calculated as below.

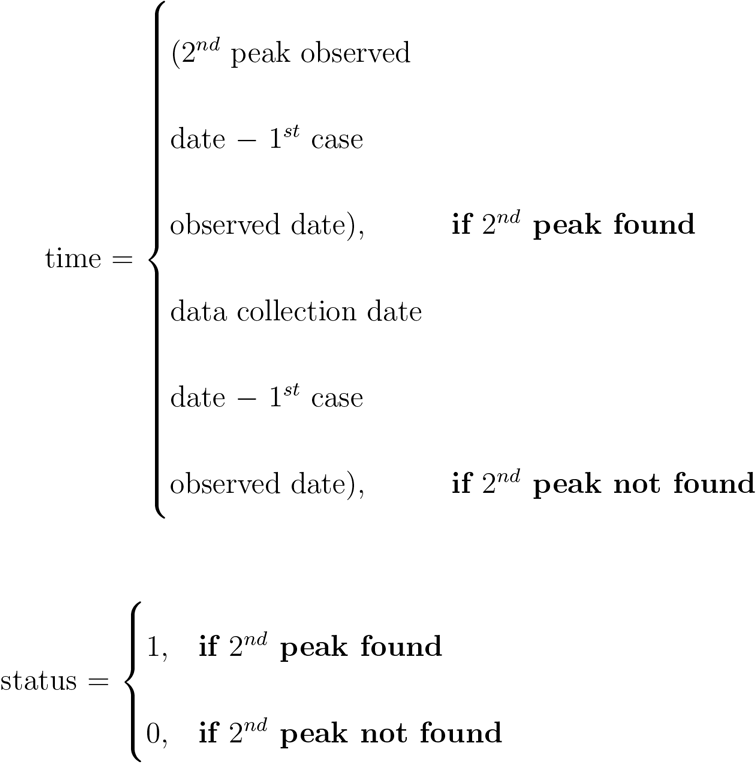

In this survival analysis study, we focussed on the *time* measured in days to hit the first and second peak. We also focussed on considering the risk assessment of factors determining these event times. The datasets were right-censored with the censoring process as defined in earlier section. All independent variables under consideration were categorical. We considered the continent, Human Development Index (HDI), population density per square killometer, GDP (Gross Deomestic Product) per capita, life expectancy (in year), percentage of people *≥* 70 years, median age (in years), diabetes prevalence in 1000, cardiovascular death rate in 100000, hospital beds per thousand, and total number of deaths all collected by January 21, 2021.

## 3 Statistical Methods

Descriptive statistics were used to describe the variables. Median of the event time to the first and second peak was calculated for different categories of the covariates. Non-parametric Kaplan–Meier survival plots were used to explore the relationship between time to reaching peak and several explanatory variables. The log-rank test was used to assess the significant difference in days to reaching peaks between the groups.

The semiparametric Cox proportional hazards (PH) model was implemented to the survival data for exploring the potential risk factors associated with the first and second peak. The Cox PH model (Cox, 1972) is essentially a regression model commonly used to model the event time data when hazard ratio is connstant between the subjects. The Cox PH model is given by

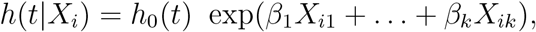

where *t* is the survival time, *h*_0_(*t*) is the baseline hazard function, *X*_*i*_ = (*X*_*i*1_, *X*_*i*2_, …, *X*_*ik*_)′ is the vector of explanatory variables and *β*= (*β*_1_,*β*_2_, …, *β*_*k*_) is the vector of regression coefficients.

The Schoenfeld residuals (Schoenfeld, 1982) were used to check the PH assumptions. We tested PH assumptions for each covariate by stating the null hypothesis that the Pearson product-moment correlation between the scaled Schoenfeld residuals and time was zero. Rejecting the null hypothesis indicated the violation of PH assumptions. All statistical analyses conducted with statistical software R.

## 4 Results

Table 1 displays characteristics of survival times over different covariates for all 188 countries used in our dataset. As of January 18, 2021, the first peak was observed in 175 countries but the second peak was observed only in 58 countries. Table 1 reveals that in the Asian and European countries the first peak occurred in quicker time than that in the countries of other continent (84 versus 109 days). Besides, the rate of happening the second peak in countries of Asia and Europe is much higher than that in the countries of other continent. In Asian and European countries, the rates of reaching the second peak were 20% higher than countries in other continents. Countries with the HDI beyond 0.85 had the fastest pace of hitting the first peak (median survival time 50 days with range 36-76), while countries with HDI of 0.65 to 0.85 had the slowest (median survival time 122.5 days with range 101-143). Furthermore, the first peak occurred at the highest rate (48.7%) in countries with a HDI of 0.85 or higher, and at the lowest rate (21.9%) in countries with a HDI of 0.65 to 0.85.

**Table 1:**
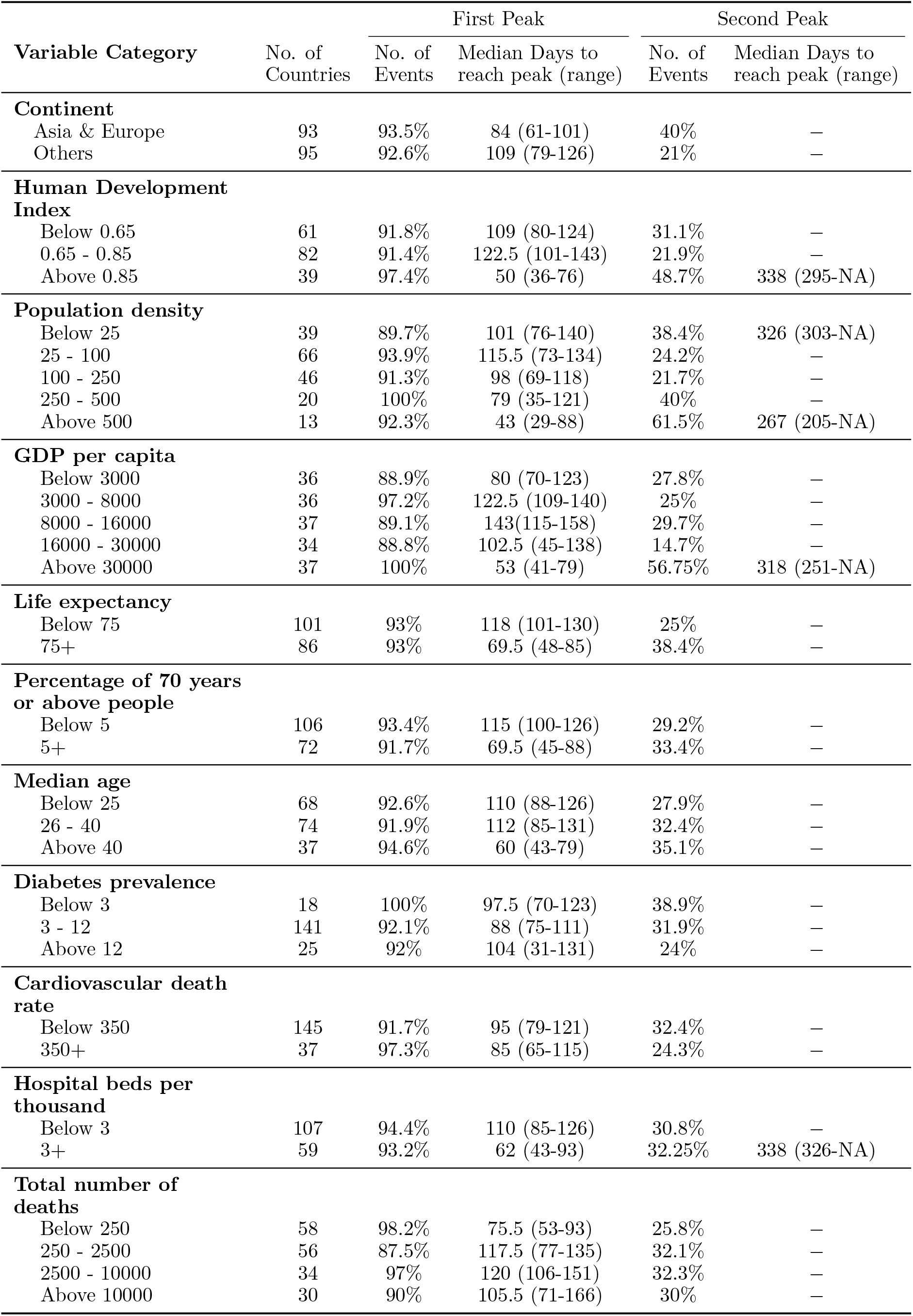
Characteristics of time to peak of global COVID-19 cases by different variables.

| Variable Category | No. of Countries | First Peak |  | Second Peak |  |
| --- | --- | --- | --- | --- | --- |
|  |  | No. of Events | Median Days to reach peak (range) | No. of Events | Median Days to reach peak (range) |
| Continent |  |  |  |  |  |
| Asia & Europe | 93 | 93.5% | 84 (61-101) | 40% | — |
| Others | 95 | 92.6% | 109 (79-126) | 21% | — |
| Human Development Index |  |  |  |  |  |
| Below 0.65 | 61 | 91.8% | 109 (80-124) | 31.1% | — |
| 0.65 - 0.85 | 82 | 91.4% | 122.5 (101-143) | 21.9% | — |
| Above 0.85 | 39 | 97.4% | 50 (36-76) | 48.7% | 338 (295-NA) |
| Population density |  |  |  |  |  |
| Below 25 | 39 | 89.7% | 101 (76-140) | 38.4% | 326 (303-NA) |
| 25 - 100 | 66 | 93.9% | 115.5 (73-134) | 24.2% | — |
| 100 - 250 | 46 | 91.3% | 98 (69-118) | 21.7% | — |
| 250 - 500 | 20 | 100% | 79 (35-121) | 40% | — |
| Above 500 | 13 | 92.3% | 43 (29-88) | 61.5% | 267 (205-NA) |
| GDP per capita |  |  |  |  |  |
| Below 3000 | 36 | 88.9% | 80 (70-123) | 27.8% | — |
| 3000 - 8000 | 36 | 97.2% | 122.5 (109-140) | 25% | — |
| 8000 - 16000 | 37 | 89.1% | 143(115-158) | 29.7% | — |
| 16000 - 30000 | 34 | 88.8% | 102.5 (45-138) | 14.7% | — |
| Above 30000 | 37 | 100% | 53 (41-79) | 56.75% | 318 (251-NA) |
| Life expectancy |  |  |  |  |  |
| Below 75 | 101 | 93% | 118 (101-130) | 25% | — |
| 75+ | 86 | 93% | 69.5 (48-85) | 38.4% | — |
| Percentage of 70 years or above people |  |  |  |  |  |
| Below 5 | 106 | 93.4% | 115 (100-126) | 29.2% | — |
| 5+ | 72 | 91.7% | 69.5 (45-88) | 33.4% | — |
| Median age |  |  |  |  |  |
| Below 25 | 68 | 92.6% | 110 (88-126) | 27.9% | — |
| 26 - 40 | 74 | 91.9% | 112 (85-131) | 32.4% | — |
| Above 40 | 37 | 94.6% | 60 (43-79) | 35.1% | — |
| Diabetes prevalence |  |  |  |  |  |
| Below 3 | 18 | 100% | 97.5 (70-123) | 38.9% | — |
| 3 - 12 | 141 | 92.1% | 88 (75-111) | 31.9% | — |
| Above 12 | 25 | 92% | 104 (31-131) | 24% | — |
| Cardiovascular death rate |  |  |  |  |  |
| Below 350 | 145 | 91.7% | 95 (79-121) | 32.4% | — |
| 350+ | 37 | 97.3% | 85 (65-115) | 24.3% | — |
| Hospital beds per thousand |  |  |  |  |  |
| Below 3 | 107 | 94.4% | 110 (85-126) | 30.8% | — |
| 3+ | 59 | 93.2% | 62 (43-93) | 32.25% | 338 (326-NA) |
| Total number of deaths |  |  |  |  |  |
| Below 250 | 58 | 98.2% | 75.5 (53-93) | 25.8% | — |
| 250 - 2500 | 56 | 87.5% | 117.5 (77-135) | 32.1% | — |
| 2500 - 10000 | 34 | 97% | 120 (106-151) | 32.3% | — |
| Above 10000 | 30 | 90% | 105.5 (71-166) | 30% | — |

The first peak had been hit by all countries with population densities of 250 to 500 square kilometers. The rates of hitting the second peak, on the other hand, were highest (61.5%) in countries with a population density greater than 500 square kilometers. Also, Countries with a population density of 25 to 100 square kilometers (median survival time 115.5 days with range 73-134) were about three times slower than countries with a population density of more than 500 square kilometers (median survival time 43 days with range 29-88) in reaching the first peak. Countries with GDP per capita over $30,000 had the highest rates of reaching both the first and second peaks (100% and 56.75%, respectively), whereas countries with GDP between $16,000 and $30.000 had the lowest percentages (88.8 & 14.7). Moreover, countries with a GDP ranging from 8000*to*16,000 had the greatest average number of waiting days (143 with a range of 115-158) to the first peak, whereas countries with a GDP above $30,000 had the shortest average number of waiting days (53 with a range of 41-79) to the first peak.

Countries with a life expectancy of fewer than 75 years experienced the first peak at a slower rate than other countries (median survival time 118 versus 69.5 days). Furthermore, the rate of the second peak happening in countries with a life expectancy of lesser than 75 years was significantly lower than in other countries (25% versus 38.4%). Countries with a fewer than 5% of the population over 70 years old (median survival time 115 days with range 100-126), a cardiovascular death rate of less than 350 (median survival time 95 days with range 79-121), and fewer than three hospital beds per thousand (median survival time 110 days with range 85-126) are slower to reach the first peak than other countries. Furthermore, The average for reaching the second peak was 338 days for countries with more than three hospital beds per thousand. Countries with a median age of more than 40 years had the fastest rate of reaching the first peak (median survival time 60 days, range 43-79), while countries with a median age of less than 25 years (median survival time 110 days, range 88-126) and countries with a median age of 26 to 40 years (median survival time 112 days, range 88-126) had nearly identical rates of reaching the first peak.

All countries with a diabetes prevalence of less than three had reached the first peak. Similarly, countries with a diabetes prevalence of less than three had the highest rates of reaching the second peak (38.9%). Also, countries with a diabetes prevalence of 3 to 12 were the quickest to reach the first peak (median survival time 88 days with a range of 75-111), whereas countries with a diabetes prevalence of more than 12 were the slowest (median survival time 104 days with a range of 31-131). The first peak occurred at the highest rate (98.2%) in countries with fewer than 250 deaths and at the lowest rate (87.5%) in countries with 250 to 2500 deaths. Additionally, countries having a total number of deaths less than 250 (median survival time 75.5 days, range 53-93) experienced the first peak 1.6 times faster than countries with a total number of deaths 2500 to 10000 (median survival time 120 days with range 106-151).

Figure 2 shows the comparison of days to reach first peak between categories of covariates by Kaplan–Meier plots. The log–rank test indicates that there was significant difference between days of reaching the first peak between group of HDI (*p* < 0.05), GDP per capita (*p* < 0.05), life expectancy (*p* < 0.05), and total number of deaths (*p* < 0.05) at 5% level of significance. However, insignificant difference in days of reaching first peak was observed between groups of continents (*p* = 0.65), population density (*p* = 0.37), percentage of people aged 70 or older (*p* = 0.46), median age (*p* = 0.39), diabetes prevalence (*p* = 0.39), cardiovascular death rate (*p* = 0.22), and hospital beds per thousand (*p* = 0.25). The Kaplan-Meier plot for countries with a life expectancy of fewer than 75 years shows that the first peak of COVID-19 cases in these countries is more likely to be longer than that for the countries with a life expectancy of 75 years or above.

**Figure 2:**
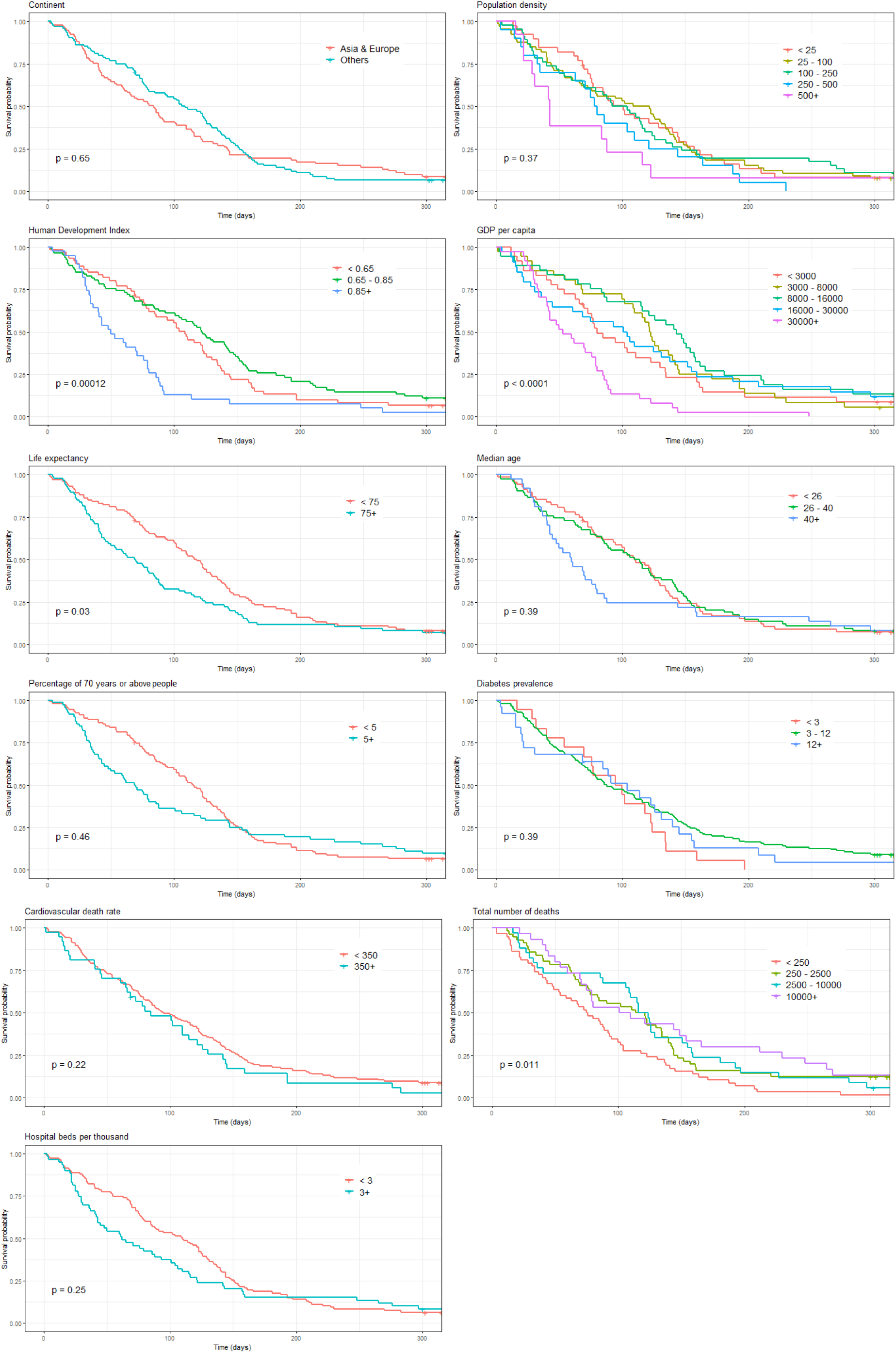
Kaplan–Meier plots for the time to getting the first peak by different covariates

Figure 3 shows that there was a significant difference in days of reaching the second peak between different group of continents (*p* < 0.05), population density (*p* < 0.05), HDI (*p* < 0.05), and GDP per capita (*p* < 0.05). But no significant difference was observed in days of reaching the second peak between categories of life expectancy (*p* = 0.096), median age (*p* = 0.93), percentage of people aged 70 or older (*p* = 0.46), total number of deaths (*p* = 0.89), diabetes prevalence (*p* = 0.39), cardiovascular death rate (*p* = 0.22), and hospital beds per thousand (*p* = 0.25). These Kaplan–Meier plots also provide clear evidence to support that there is significance difference in the numbers of days to reach first and second peaks over different categories of the variables considered in this study.

**Figure 3:**
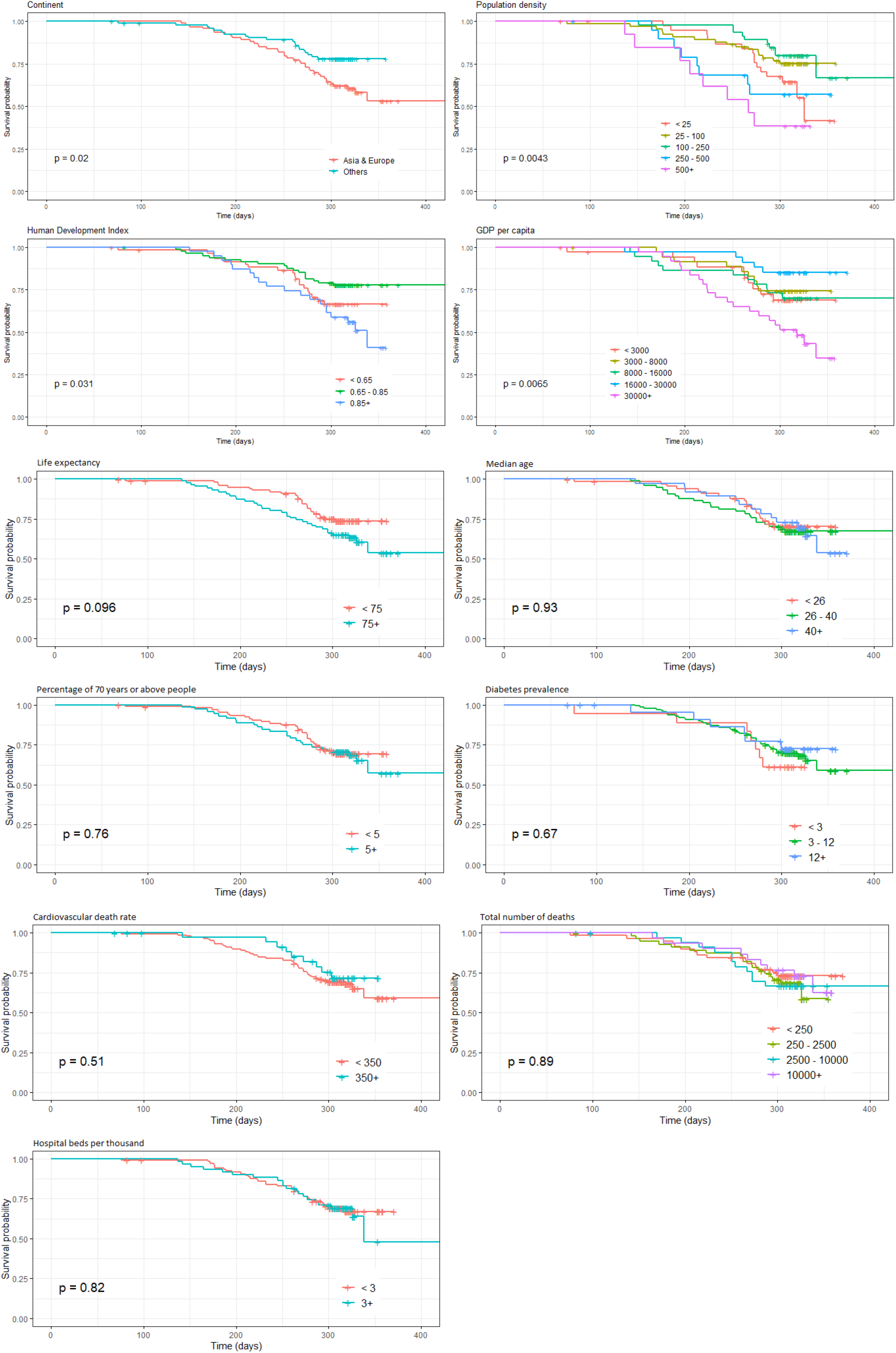
Kaplan–Meier plots for the time to getting the second peak by different covariates

Table 3 suggests that the implementation of the Cox PH model with the covariates satisfies all basic assumptions of the model. Before implementing the Cox PH model, these assumptiones were checked as part of the model diagnosis. Results of the Cox PH model (Table 4) show that compared to countries under “other” continents, Asian & European conitent countries has 75% more chance of hitting the first peak of COVID-19 cases. On the contrary, contrast results are found for the case of second peak. Statistically insignificant difference between the chance of hitting the second peak across categories of the variable continent was found.

**Table 3:** Product-moment correlation between schoenfeld residual vs time to COVID-19 peak.

| Variable Category | First Peak |  |  | Second Peak |  |  |
| --- | --- | --- | --- | --- | --- | --- |
| | $\rho$ | $\chi^2$ | $p$ value | $\rho$ | $\chi^2$ | $p$ value |
| **Continent** |  |  |  |  |  |  |
| Asia & Europe | 0.075 | 1.008 | 0.315 | -0.258 | 5.928 | 0.015 |
| **Human Development Index** |  |  |  |  |  |  |
| 0.65 - 0.85 | 0.014 | 0.284 | 0.594 | 0.075 | 0.293 | 0.589 |
| Above 0.85 | -0.022 | 0.264 | 0.607 | 0.110 | 1.081 | 0.299 |
| **Population density** |  |  |  |  |  |  |
| 25 - 100 | 0.058 | 0.004 | 0.952 | -0.178 | 0.394 | 0.530 |
| 100 - 250 | -0.011 | 0.704 | 0.401 | -0.037 | 0.721 | 0.396 |
| 250 - 500 | 0.053 | 0.685 | 0.408 | -0.287 | 2.122 | 0.145 |
| Above 500 | -0.037 | 1.233 | 0.267 | -0.166 | 0.962 | 0.327 |
| **GDP Per capita** |  |  |  |  |  |  |
| 3000 - 8000 | 0.197 | 5.939 | 0.015 | -0.195 | 1.453 | 0.228 |
| 8000 - 16000 | 0.149 | 0.336 | 0.562 | -0.022 | 0.280 | 0.596 |
| 16000 - 30000 | 0.116 | 9.704 | 0.002 | -0.026 | 0.054 | 0.816 |
| Above 30000 | 0.237 | 1.146 | 0.284 | -0.046 | 1.263 | 0.261 |
| **Life expectancy** |  |  |  |  |  |  |
| 75+ | -0.085 | 4.872 | 0.027 | -0.120 | 0.198 | 0.656 |
| **Percentage of 70 years or above people** |  |  |  |  |  |  |
| 5+ | -0.242 | 19.018 | 0.000 | -0.230 | 0.924 | 0.336 |
| **Median age** |  |  |  |  |  |  |
| 26 - 40 | -0.032 | 0.071 | 0.790 | 0.162 | 1.761 | 0.184 |
| Above 40 | 0.004 | 2.756 | 0.097 | 0.323 | 3.524 | 0.060 |
| **Diabetes prevalence** |  |  |  |  |  |  |
| 3 - 12 | -0.092 | 0.618 | 0.432 | -0.111 | 0.017 | 0.896 |
| Above 12 | -0.112 | 0.140 | 0.709 | -0.188 | 0.072 | 0.788 |
| **Cardiovascular death rate** |  |  |  |  |  |  |
| 350+ | -0.028 | 0.119 | 0.730 | 0.192 | 3.457 | 0.063 |
| **Hospital beds per thousand** |  |  |  |  |  |  |
| 3+ | -0.011 | 6.824 | 0.009 | -0.264 | 0.035 | 0.852 |
| **Total number of deaths** |  |  |  |  |  |  |
| 250 - 2500 | 0.081 | 0.188 | 0.664 | 0.159 | 0.949 | 0.330 |
| 2500 - 10000 | 0.104 | 0.672 | 0.412 | 0.126 | 0.401 | 0.526 |
| Above 10000 | 0.066 | 0.037 | 0.847 | 0.071 | 0.157 | 0.692 |

**Table 4:** Cox proportional hazard estimates and their 95% confidence intervals.

| Variable Category | First Peak |  | Second Peak |  |
| --- | --- | --- | --- | --- |
|  | Hazard Ratio | 95% Confidence Interval | Hazard Ratio | 95% Confidence Interval |
| **Continent** |  |  |  |  |
| Others | 1 |  | 1 |  |
| Asia & Europe | 1.753* | (1.046, 2.936) | 0.649 | (0.227, 1.855) |
| **Human development index** |  |  |  |  |
| Below 0.65 | 1 |  | 1 |  |
| 0.65 - 0.85 | 0.349* | (0.148, 0.820) | 0.085* | (0.008, 0.915) |
| Above 0.85 | 0.572 | (0.172, 0.920) | 0.030* | (0.002, 0.584) |
| **Population density** |  |  |  |  |
| Below 25 | 1 |  | 1 |  |
| 25 - 100 | 1.182 | (0.689, 2.027) | 0.538 | (0.210, 1.379) |
| 100 - 250 | 0.965 | (0.555, 1.680) | 0.329* | (0.119, 0.908) |
| 250 - 500 | 2.259* | (1.149, 4.444) | 1.805 | (0.617, 5.278) |
| Above 500 | 1.294 | (0.488, 3.249) | 1.795 | (0.496, 6.497) |
| **Life expectancy** |  |  |  |  |
| Below 75 | 1 |  | 1 |  |
| 75+ | 1.871* | (1.003, 3.490) | 3.1 | (0.745, 12.899) |
| **GDP per capita** |  |  |  |  |
| Below 3000 | 1 |  | 1 |  |
| 3000–8000 | 1.422 | (0.650, 3.108) | 0.498 | (0.082, 3.039) |
| 8000–16000 | 2.303 | (0.798, 6.643) | 0.2514 | (0.161, 39.311) |
| 16000–30000 | 2.416 | (0.858, 6.797) | 1.373 | (0.084, 22.424) |
| Above 30000 | 9.579* | (2.436, 37.658) | 15.085 | (0.565, 400.830) |
| **Percentage of 70 years or above people** |  |  |  |  |
| Below 5 | 1 |  | 1 |  |
| 5+ | 0.742 | (0.411, 1.339) | 0.861 | (0.246, 3.016) |
| **Median age** |  |  |  |  |
| Below 26 |  |  | 1 |  |
| 26 - 40 | 1.004 | (0.498, 2.023) | 2.281 | (0.361, 14.434) |
| Above 40 | 0.600 | (0.208, 1.729) | 1.017 | (0.113, 9.145) |
| **Diabetes prevalence** |  |  |  |  |
| Below 3 | 1 |  | 1 |  |
| 3 - 12 | 0.451 | (0.201, 1.010) | 0.155* | (0.036, 0.680) |
| Above 12 | 0.392 | (0.138, 1.112) | 0.113* | (0.014, 0.898) |
| **Cardiovascular death rate** |  |  |  |  |
| Below 350 | 1 |  | 1 |  |
| 350+ | 2.941*** | (1.595, 5.426) | 1.497 | (0.430, 5.213) |
| **Hospital beds Per thousand** |  |  |  |  |
| Below 3 | 1 |  | 1 |  |
| 3+ | 1.902* | (1.099, 3.291) | 1.680 | (0.646, 4.371) |
| **Total number of deaths** |  |  |  |  |
| Below 250 | 1 |  | 1 |  |
| 250 - 2500 | 0.595* | (0.370, 0.958) | 0.690 | (0.286, 1.665) |
| 2500 - 10000 | 0.654 | (0.374, 1.143) | 1.862 | (0.716, 4.84) |
| Above 10000 | 0.402** | (0.223, 0.722) | 0.860 | (0.304, 2.428) |

\* $p$ value < 0.05; \*\* $p$ value < 0.01; \*\*\* $p$ value < 0.001

Countries which have HDI between 0.65 and 0.85 are 65% less likely to experience the first peak of COVID-19 cases than the countries which have HDI below 0.65. But no significant difference in the chances of hitting the first peak between countries with HDI below 0.65 and above 0.85. Countries which have HDI between 0.65 and 0.85 and above 0.85 are 92.5% and 97% less likely to experience the second peak of COVID-19 cases respectively than the countries with HDI below 0.65.

Only the category ‘250-500’ of variable population density has a significant hazard ratio (*p* value < 0.05) with the first peak. Countries with a population density, 250 to 500 are 2.25 times more likely to experience the first peak of COVID-19 cases than the countries with a population density below 25. The category ‘100 -250’ is significant with the second peak. Countries with population density, 100 to 250 are 67% less likely to experience COVID-19 second peak than the countries with a population density below 25.

Life expectancy is a significant factor for getting only the first peak but the second peak of COVID-19 cases. Countries which have life expectancy above 75 years are almost 87% more likely to get the first peak compared to countries which having life expectancy below 75 years. The variable GDP per capita holds only one category (Above USD 30000), which had a significant hazard ratio with the first peak. Countries with GDP per capita above USD 30000 are 9.5 times likely to experience the first peak of COVID-19 cases than countries with GDP below USD 3000. Diabetes prevalence is the significant factor for the second peak. Countries with diabetes prevalence, 3 to 12 and above 12 are 84% and 89% less likely to experience the second peak of COVID-19 cases respectively both compared to the countries which have less than 3 diabetes prevalence.

Countries which have the cardiovascular death rate above 350 per 100000 people are almost 2.94 times more likely to get the first peak compared to the countries which have cardiovascular death rate below 350 per 100000 people. There was no significant difference between the chances of hitting the second peak across the categories of variable hospital beds per thousand. Countries which have hospital beds above 3 per thousand people has 1.9 times more chance to experience the first peak of COVID-19 cases compared to the countries which have hospital beds less than 3. The hazard ratio for the countries with a total number of deaths 250 to 2500’ and ‘Above 10000’ is 0.595 and 0.402 respectively. Countries which have a total number of deaths between 250 and 2500, and above 10000 are 41% and 60% respectively more likely to experience the COVID-19 first peak than countries which has a total number of deaths below 250.

## 5 Discussions and Conclusions

This paper sheds light on the awareness of factors associated with the COVID-19 first and second waves that are mentioned exchangebly in this study as the first and second peak respectively. We investigated the median time, measured in days, of hitting the first and second peak of COVID-19 cases for 188 countries respective to the presence of several covariates. We gave the defination of the first and second peak on the basis of data-driven evidence for respective country. We identified a number of covariates that has a substantial impact on number of days of hitting the peaks. We found that the median number of days to the first peak was considerably small in Asian & European countries compared to other continents, in countries where HDI is above 0.85, in countries where population density is higher (e.g., *>* 250 per sq. km), in countries where GDP per capita is either below USD 3000 or above 30000, in countries where life expectancy is above 75, in countries where median age is above 40, in countries where diabetes prevalence is between 3 and 12, in countries where cardiovascular death rate is above 350 per 100000, in countries where number of hospital beds is above 3 per 1000.

The second peak was observed for the countries where some of the covariates including HDI, population density, GDP per capita and hospital beds played vital role. The median number of days to hit second peak was observed as 338 days for the countries where HDI is above 0.85, as 326 and 267 days for the countries where population density is below 25 and above 500 respectively, as 318 days for the countries where GDP per capita is above USD 30000, and as 338 days for the countries where hospital beds per thousand is above 3.

Results revealed that the percentage of second peak is substantially higher in Asian & European continent countries. However, it appeared that the second peak was more likely to occur in countries where population densely is higher, GDP per capita is higher, life expectancy is higher, median age is higher, diabetes prevalence is lower, cardiovascular death rate is below 350, and hospital beds per thousand is above 3.

Fitting the multivariable Cox proportional hazards regression model for the days to reach both the first and second peak also reveals that severral factors including the continent, Human Development Index, population density, GDP per capita, life expectancy, cardiovascular death rate, hospital beds per thousand, and total number of deaths are significantly associated with the first peak of COVID-19 cases. However, only three factors– Human Development Index, population density, and diabetes prevalence are found as significant factors that are associated with the second peak of COVID-19 cases for the countries.

More specifically we found that countries having HDI between 0.65 and 0.85 are 65% and 92.5% less likely to experience the first peak and second peak of COVID-19 cases respectively than the countries having HDI below 0.65. Countries having population density between 250 and 500 are 2.25 times more likely to experience the first peak of COVID-19 cases than the countries having population density below 25. Countries having GDP per capita above USD 30000 are 9.5 times likely to experience the first peak of COVID-19 cases than countries having GDP below USD 3000. Countries having the cardiovascular death rate above 350 per 100000 people are almost 2.94 times more likely to get the first peak compared to the countries having cardiovascular death rate below 350 per 100000 people.

This study has some limitations. The data used contained several missing values that were omitted before performing the Cox PH model. We did not apply any missing value technique to impute them. Some essential variables including hand washing facilities, smokers prevalence, hospital patients per million, poverty rate, etc. have not been considered in the analysis due to data deficiency.

## Data Availability

The working data set used for this study could be sent upon request.

## Competing Interests

We declare that we have no competing interests.

## Conflict of interest

The authors declare no conflict of interest.

## Funding

There is no funding for this study.

## Author’s Contributions

YAO contributed to designing, analysing and drafting the manuscript, both BZ and DB contributed to analysing and finalizing the drafting, MHRK contributed to designing methods and manuscript, arranging data, and primary drafting.

## Availability of Data and Materials

The working data set used for this study has been submitted to the journal as additional supporting file.

## Notes

### Competing Interest Statement

The authors have declared no competing interest.

